# Utility of *R*_t_ epidemic trend categories for influenza surveillance: alignment with observed emergency department trends across two influenza seasons in the United States

**DOI:** 10.64898/2026.11.02.26367103

**Authors:** Sarabeth M. Mathis, Rebecca K. Borchering, Zachary Susswein, Nate McIntosh, Patrick Corbett, Kelly Carey, Alexia Couture, Krista Kniss, Katie Reinhart, Daneisha Hawkins, Sarah Connolly, Danielle Richard, Adam Howes, Alicia Budd, Katelyn Gostic, Matthew Biggerstaff

## Abstract

CDC has estimated the time-varying reproduction number (*R*_t_) for influenza in the United States since 2023, translating the posterior probability that *R*_t_ exceeds one, *P(R*t *> 1)*, into epidemic trend categories (growing, likely growing, not changing, likely declining, declining). If *R*_t_ reliably indicates infection dynamics, growing or declining trends should precede corresponding changes in influenza-related emergency department (ED) visits, accounting for the lag between infection and care-seeking. However, because these categories reflect confidence in the direction of change rather than magnitude, their alignment with the corresponding trends ED activity measures is unclear.

We compared weekly *R*_t_ epidemic trend categories to observed ED trends from the National Syndromic Surveillance Program across the 2024/2025 and 2025/2026 influenza seasons, using three thresholds to define observed trends at different magnitudes (≥0.1% difference, ≥0.5% difference, ≥5% relative change between consecutive weeks), comparing *R*_t_ trends to observed trends the following week. We assessed categorical alignment and calibration between *P(R*t *> 1*) and observed trends.

Alignment was moderate overall, strongest for growing and declining categories under the 5% relative change threshold (61% aligned) and weakest under the 0.5% difference threshold (38.2% aligned); calibration was similarly best under the 5% relative change threshold. Misalignment concentrated around epidemic inflection points and periods of low influenza activity.

*R*_t_ epidemic trend categories showed moderate, threshold-dependent alignment with observed ED activity, reflecting a combination of real-time model error and misalignment between direction-based and magnitude-based surveillance metrics. Defining categories based on *R*_t_ magnitude could improve correspondence with size-based change indicators and strengthen their utility for real-time monitoring.

## Introduction

Influenza remains a significant public health concern, with seasonal outbreaks leading to substantial morbidity and mortality in the US[1]. Individuals as well as public health and healthcare partners benefit from real-time situational awareness including influenza surveillance summaries, forecasts, and nowcasts. One such metric is the time-varying reproduction number (*R*_t_) which estimates the average number of new infections that each infected person causes at a specific time during a disease epidemic. *R*_t_ implicitly reflects population susceptibility, contact patterns, and virus transmissibility[2, 3]. Unlike static measures of transmissibility such as R_0_, *R*_t_ can provide timely insights into whether an outbreak is growing or declining. These could help public health practitioners, healthcare systems, and frontline responders with routine situational awareness, response readiness, and resource planning.

CDC has been estimating the effective reproduction number for influenza and COVID-19 since 2023, and RSV since October 2025, to help stakeholders monitor respiratory viruses and plan for potential increases in disease activity in combination with other surveillance metrics. CDC developed five categories (growing, likely growing, not changing, likely declining, and declining) to summarize the likelihood the epidemic trend is classified as growing based on the posterior probability, *P(R*t *> 1)*[4].

In an idealized system where *R*_t_ is known, rising or falling infections on date *t* (*R*_t_ >1 or *R*_t_ < 1) will precede measurable changes in influenza-associated emergency department (ED) visits (accounting for lags between infection and care seeking) under the following conditions: if *R*_t_ is approximately constant from the date of infection *(t)* to the date of ED visit (*t + d),* and if the rate of change in infections, (and in turn, in ED visits) indicated by *R_t_,* exceeds the detection threshold for changes in ED visits. In practice, misalignment may occur if *R_t_* is misestimated, or if it changes abruptly.

Another potential source of misalignment is differences in how increasing and decreasing trend categories are defined for public reporting of ED visits and *R_t_*. Specifically, observed increases occur when the rate or count of new ED visits, positive tests, or similar measures rise beyond a specified threshold of change. Meanwhile, CDC currently defines increasing transmission trends based on the posterior probability *P(R*t *> 1)*, which indicates confidence that infections are increasing, but provides no information about the expected magnitude of change.

Taking these potential sources of misalignment into account, this evaluation assesses whether reported growing or declining infection trend categories, based on *R_t_* estimates, preceded corresponding changes in ED visits, with some expected lag between infection and care-seeking. To address this, we evaluated *R*_t_ epidemic trend categories as early indicators of changing influenza ED activity by assessing their alignment with subsequent observed trends in influenza ED visits. This analysis evaluates the extent to which direction-based *R_t_* epidemic trend categories correspond to observed changes in ED activity. It also identifies how differences in the definitions of these surveillance metrics may affect their alignment.

## Methods

We compared the *R*_t_ epidemic trends from the 2024/2025 and 2025/2026 influenza seasons to weekly changes in the percentage of influenza-diagnosed ED visits from the National Syndromic Surveillance Program (NSSP) using three different definitions, used by CDC, of a meaningful increase or decrease in ED activity.

### Categorizing Epidemic Trends from R_t_ Data

CDC estimated *R*_t_ for the US overall, each state with available data, and Washington DC using daily counts of ED visits diagnosed with influenza from NSSP. Detailed methods for determining *R*_t_ have been described fully elsewhere[3, 5]. Briefly, CDC fit renewal models to real-time influenza ED visit data using EpiNow2, which included nowcasting adjustments for incomplete reporting on the most recent dates [3, 6]. *R*_t_ estimates were converted into a categorical indicator based on the proportion of the posterior probability greater than one, which would indicate that the epidemic was growing[4]. This metric, referred to as the *R*_t_ epidemic trend, summarized the model’s posterior probability that infections were growing, not changing, or declining based on the proportion of the distribution above one:

- Growing: >90% of the distribution of *R*_t_ >1
- Likely growing: 76%-90% of the distribution of *R*_t_ >1
- Not Changing: 26%-75% of the distribution of *R*_t_ >1
- Likely declining: 10%-25% of the distribution of *R*_t_ >1
- Declining: <10% of the distribution of *R*_t_ >1
- Not estimated: *R*_t_ estimates were not published

*R*_t_ was not published for a jurisdiction when data quality standards were not met, including insufficient ED visit volume, detected reporting anomalies, or model reliability checks that were not passed. *R*_t_ estimates and epidemic trends were published on Friday each week during the influenza season [7]. Weeks were considered not estimated if the *R*_t_ estimate was not published, even though an unreliable estimate may have been available. Weeks where *R*_t_ was not estimated were included in tables and figures shown but excluded from alignment analyses.

In a typical week, models to estimate *R*_t_ were run on Wednesday morning, using real-time NSSP data. The *R*_t_ trend estimate published each week reflected data as of the most recent Tuesday. *R*_t_ estimates and epidemic trend categories were reviewed each week by modelers, pathogen subject matter experts, and data stewards.

The *R*_t_ estimation pipeline used daily counts of ED visits from a fixed set of facilities active as of 2023, meaning the contributing facilities did not change across the course of the *R*_t_ estimation period. However, these data required manual review to identify and address unplanned interruptions or changes in reporting, as the *R*_t_ estimation method was sensitive to changes in the fraction of infections observed as ED visits.

### Categorizing Observed Trends from NSSP Data

For validation, we compared predicted real-time *R*_t_ estimates to observed weekly changes in the percent and count of NSSP ED visits with a discharge diagnosis of influenza, which is a publicly reported signal used to monitor influenza activity at the federal and jurisdictional levels [8–10]. We used these data published July 3, 2026. From this dataset, weekly percentages were reported Friday and include data through the end of the most recent MMWR week (the preceding Saturday) encompassing all emergency department visits. Reporting anomalies in real-time provisional data are often corrected over time and would have been updated in these data, but unlike the dataset that was used for *R*_t_ estimation, this dataset did not adjust for onboarding of new facilities or interruptions in data transmission; however, it did allow for onboarding, drop-outs, and re-onboarding of facilities.

For each jurisdiction during the 2024/2025 and 2025/2026 influenza seasons, we calculated the week-to-week differences in the observed percentage of weekly ED visits with a discharge diagnosis for influenza and week-to-week relative changes in observed number of influenza ED visits. Total ED visits are reasonably stable over time. Therefore, the observed percent of influenza ED visits can serve as proxies for normalized incidence, differences can be interpreted as indicators of absolute changes in incidence, and relative change can be interpreted as an indicator of epidemic growth rate. Calculations were between week *w* (the MMWR week in which the *R*_t_ trend estimate was generated) and the following week, *w + 1*, which was unobserved when the *R*_t_ estimate was made for each jurisdiction (Supplement Figure 1).

From these differences and relative changes, we defined three ED trend categories (increase, decrease, stable) using three thresholds:

- Increase: a positive absolute difference of ≥0.1%, ≥0.5%, or >5% relative change
- Decrease: a negative absolute difference of ≤-0.1%, ≤-0.5%, or <-5% relative change
- Stable: any absolute difference between -0.1% – 0.1%, -0.5% – 0.5%, or -5% – 5% relative change

All three thresholds are used in CDC’s FluView report to determine whether influenza activity has changed week-to-week in different surveillance systems [9, 11]. The 0.1% difference threshold is used to detect changes in the percentage of outpatient visits with an influenza-like-illness, and the 0.5% difference threshold is used to detect changes in the percentage of specimens testing positive for influenza in clinical laboratories. The 5% relative change threshold was used to detect changes in the number of weekly influenza-related hospital admissions during the 2024-2025 season. Relative change metrics have a direct relationship to *R*_t_ (*R*_t_ is a transformation of the epidemic growth rate and can be interpreted as the per-generation rate of epidemic growth)[12]. Difference metrics indicate changes in absolute incidence, and consequently are insensitive when influenza incidence is low. For example, it is not possible to detect a 0.1% increase in ED visits while the percentage of influenza ED visits remains below 0.1%. Changes in infection or ED visit incidence depend on the current epidemic growth rate (measured by *R*_t_) and on current epidemic size. Thus, they have an indirect relationship to *R*_t_, and this relationship changes over the course of the influenza season as epidemic size rises and falls. Relative change may more closely reflect the proportional changes captured by *R*_t_ at any point in the season.

### Data alignment

*R*_t_ epidemic trend category estimates and ED trend category data were aligned by MMWR week (*w*) and jurisdiction. Wyoming and Missouri were excluded from the 2024/2025 and a portion of the 2025/2026 season due to insufficient or unavailable NSSP data during that period. Once a reliable data feed was established during the 2025/2026 season, CDC began publishing epidemic trend estimates for each state.

Given an expected lag of approximately one week between infection and ED presentation, accounting for the influenza incubation period of approximately two days (range 1–4 days) [13] and an additional delay from symptom onset to care-seeking, a growing epidemic trend estimated from Tuesday data (week *w*) would be expected to correlate with increases in ED visits within the following MMWR week, which ends the subsequent Saturday (week *w + 1*).

### Assessment of alignment

For each jurisdiction-week, we assessed alignment of categories between the *R*_t_ epidemic trend and observed ED trend categories. Trends were considered aligned when both indicated change in the same direction (growing or likely growing with increasing, or declining or likely declining with decreasing) or when both indicated stability (not changing with stable). Major misalignments were defined as changes in opposite direction: growing or likely growing with decreasing, or declining or likely declining with increasing. All remaining combinations, where one source indicated stability while the other indicated directionality, were classified as misalignments. We also examined alignment across each season by state and week to assess whether alignment patterns varied geographically or temporally.

We also assessed whether the probability that *R*_t_ was growing, the metric underlying the epidemic trend categories, corresponded to the frequency of observed increases in ED visits. We did not expect a direct, linear relationship between predicted probability and the magnitude of observed change since the probability metric reflects confidence in the direction of the epidemic trend rather than its rate or size of change. Therefore, we evaluated calibration rather than correlation by binning the predicted probability of increasing infections into 0.1-wide bins *R*_t_ from 0 to 1, and within each bin calculated the observed proportion of jurisdiction-weeks with an observed increase in subsequent ED visits for each threshold. If the predicted probabilities were well-calibrated, the observed proportion of increases in each bin should fall close to the 1:1 line (i.e., a bin with predicted probability of growth near 0.9 should show approximately 90% of jurisdiction-weeks with an observed increase).

To illustrate the underlying relationship between the probability that *R*_t_ was growing and the magnitude of observed change – and the conceptual distinction between direction-based and magnitude-based trend definitions – we additionally visualized the probability that *R*_t_ was growing against the two continuous ED visit-based metrics underlying the trend category thresholds: the week-to-week absolute difference in the percentage of ED visits due to influenza and the week-to-week relative percent change in the number of ED visits due to influenza. Given that the predicted probability reflects confidence in direction rather than magnitude, this relationship is not expected to be linear or consistently monotonic. A predicted probability near 0.5 corresponds to median *R*_t_ estimates near 1, indicating an underlying growth rate close to zero; we therefore anticipated that observed changes in ED visits would be small in magnitude when predicted probability was near 0.5. As predicted probability moved toward 0 or 1, this constraint relaxes, since *R*_t_ confidently above or below 1 is compatible with a wide range of growth or decline rates. We therefore anticipated a wider range of observed changes, including larger-magnitude increases or decreases, at the extremes of the probability distribution, without expecting every jurisdiction-week at high or low probability to show a large change.

### Sensitivity analysis

As a sensitivity analysis, we repeated all analyses using weeks *w— 1* and *w* to define the observed ED trend, where *w* was the MMWR week in which the *R*_t_ trend estimate was generated and *w— 1* was the preceding week, which was observed at the time the estimate was made. This allowed us to assess whether *R*_t_ epidemic trend categories performed differently when compared to concurrent observed trends rather than trends in the immediate following week, testing whether *R*_t_ functions as an early indicator rather than a concurrent one. Results from this analysis are available in the Supplement.

Daily counts of influenza-diagnosed ED visits used to estimate *R_t,_*and weekly counts used to establish the 5% relative change threshold are not uniformly publicly available. Weekly percentages of ED visits with a discharge diagnosis for influenza and *R_t_*data including epidemic trend categories are publicly available[7, 10]. All analyses were performed in R version 4.5.0.

## Results

*R*_t_ epidemic trend and observed ED trend categories were estimated for 50 jurisdictions, including the US, Washington DC, and 48 states, across 33 weeks, from October 5, 2024, through May 17, 2025, for the 2024/2025 influenza season. Missouri and Wyoming were excluded from the 2024/2025 season due to insufficient or unavailable data. For the 2025/2026 season, *R*_t_ epidemic trend and observed ED trend categories were estimated for 52 jurisdictions, including the US, Washington DC and 50 states, across 34 weeks from October 4, 2025, through May 23, 2026. Across both seasons a total of 2,976 jurisdiction-weeks were available for analysis after excluding weeks where *R*_t_ was not estimated.

Nationally, the percentage of ED visits due to influenza during the 2024/2025 season increased rapidly from approximately 1% in early December 2024 to 5.5% by late December, briefly declining to 4.5% in early January before reaching a season peak of 8.4% in early February 2025. The percentage then decreased steadily, falling below 1% by mid-April 2025. During the 2025/2026 season, the percentage increased from approximately 1% in late November 2025 to a season peak of 8.6% by late December. Following the peak, the percentage declined to below 4% by mid-January 2026 before stabilizing, fluctuating between approximately 2.5% and 4% through early March 2026. As in the prior season, the percentage fell below 1% by mid-April 2026. Figure 1 shows the percentage of ED visits for influenza nationally by week, with colored categorizations for the *R*_t_ epidemic trend for a given week, and the ED trend based on the three thresholds for a given week (0.1% difference, 0.5% difference, and 5% relative change) displayed for both seasons. Supplement Figure 2 shows the distribution of weekly absolute and relative changes across both seasons.

**Figure 1:**
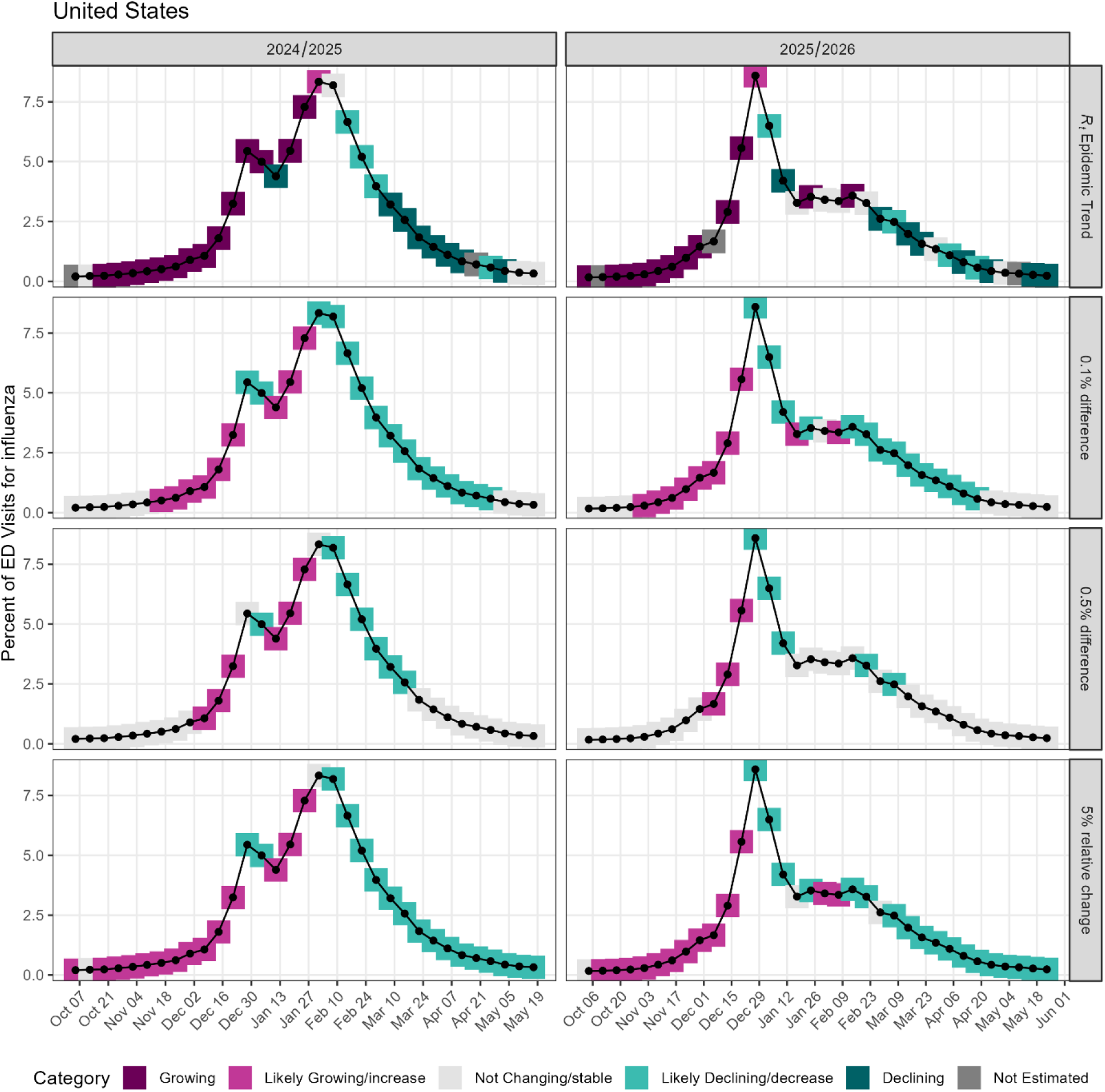
Percentage of ED visits for influenza in the US overall for the 2024/2025 and 2025/2026 seasons with boxes colored by categories defined by the *R*_t_ epidemic trend and the three ED trends (0.1% difference, 0.5% difference, and 5% relative change). The legend shows the category colors for *R_t_* epidemic trends and observed trend categories listing epidemic trend category/observed category. Observed thresholds are based on differences between week *w* and week *w + 1*.

### *R*_t_ Epidemic and ED trend alignment

Table 1 shows the distribution of *R*_t_ epidemic trend categories and observed ED trend categories across all 3,484 jurisdiction-weeks, including not estimated weeks, as well as the alignment between the two for each threshold. Declining was the most common *R*_t_ epidemic trend category (n=938, 26.9%), followed by growing (n=789, 22.6%) and not changing (n=650, 18.7%). Likely growing and likely declining were less common (n=270, 7.7% and n=329, 9.4%, respectively), and 508 jurisdiction-weeks (14.6%) had no *R*_t_ estimate and were excluded from analysis. The number of not estimated jurisdiction-weeks includes counts of Missouri and Wyoming weeks where data were unavailable during the 2024/2025 season.

**Table 1:** Count and percentage of *R*_t_ epidemic trend categories (rows) that aligned with observed ED trends defined at the 0.1% difference, 0.5% difference, and 5% relative change thresholds combined across the 2024/2025 and 2025/2026 influenza seasons. Cells with major misalignment (e.g. growing *R*_t_ epidemic trend but observed decrease in the ED trend) are bolded for visibility. Percentages in the Increase, Stable, and Decrease columns are row percentages. Observed thresholds are based on differences between week *w* and week *w + 1*.

|  | Total | Increase | Stable | Decrease |
|---|---|---|---|---|
| **Threshold 0.1% difference** | n = 3484 | n = 1003 | n = 1143 | n = 1338 |
| Growing | 789 | 551 (69.8%) | 112 (14.2%) | **126 (16%)** |
| Likely Growing | 270 | 84 (31.1%) | 116 (43%) | **70 (25.9%)** |
| Not Changing | 650 | 142 (21.8%) | 241 (37.1%) | 267 (41.1%) |
| Likely Declining | 329 | **35 (10.6%)** | 114 (34.7%) | 180 (54.7%) |
| Declining | 938 | **77 (8.2%)** | 272 (29%) | 589 (62.8%) |
| Not Estimated | 508 | 114 (22.4%) | 288 (56.7%) | 106 (20.9%) |
| **Threshold 0.5% difference** | n = 3484 | n = 531 | n = 2317 | n = 636 |
| Growing | 789 | 343 (43.5%) | 372 (47.1%) | **74 (9.4%)** |
| Likely Growing | 270 | 32 (11.9%) | 194 (71.9%) | **44 (16.3%)** |
| Not Changing | 650 | 59 (9.1%) | 436 (67.1%) | 155 (23.8%) |
| Likely Declining | 329 | **18 (5.5%)** | 219 (66.6%) | 92 (28%) |
| Declining | 938 | **35 (3.7%)** | 671 (71.5%) | 232 (24.7%) |
| Not Estimated | 508 | 44 (8.7%) | 425 (83.7%) | 39 (7.7%) |
| **Threshold 5% relative change** | n = 3482* | n = 1432 | n = 333 | n = 1717 |
| Growing | 789 | 605 (76.7%) | 58 (7.4%) | **126 (16%)** |
| Likely Growing | 270 | 151 (55.9%) | 35 (13%) | **84 (31.1%)** |
| Not Changing | 650 | 258 (39.7%) | 73 (11.2%) | 319 (49.1%) |
| Likely Declining | 329 | **64 (19.5%)** | 39 (11.9%) | 226 (68.7%) |
| Declining | 938 | **109 (11.6%)** | 70 (7.5%) | 759 (80.9%) |
| Not Estimated | 506 | 245 (48.4%) | 58 (11.5%) | 203 (40.1%) |

\*Multiple consecutive weeks for one jurisdiction had zero ED visits which made calculating relative change impossible, these weeks were classified as Not Estimated for R<sub>*t*</sub> epidemic trends and not included in the alignment analysis.

The frequency of observed ED trend categories varied considerably across thresholds. The 5% relative change threshold classified the largest proportion of jurisdiction-weeks as changing (decreasing: 49.3%, increasing 41.1%), while the 0.5% difference threshold classified the majority as stable (66.5%). The 0.1% difference threshold produced a more balanced distribution among increasing (28.8%), decreasing (38.4%) weeks, and stable (32.8%).

Agreement was strongest for growing and declining *R*_t_ categories, particularly under the 0.1% difference and 5% relative change thresholds, where 69.8% and 76.7% of growing weeks were classified as increasing and 62.8% and 80.9% of declining weeks as decreasing, respectively. The 0.5% difference threshold showed weaker alignment, with most jurisdiction-weeks classified as stable regardless of *Rt* trend category. Likely growing, likely declining, and not changing *R_t_* trend categories showed weaker alignment across all thresholds, and not changing *R_t_* trend weeks frequently had observed ED trend increases or decreases based on the 0.1% and 5% thresholds.

Table 2 shows alignment by threshold and season. Overall alignment was consistent across both seasons, ranging from 36.0% at the 0.5% difference threshold to 62.6% at the 5% relative change threshold. Major misalignment was uncommon across all thresholds and seasons, ranging from 3.0% to 7.9% of jurisdiction-weeks, and was highest for the 5% relative change threshold and lowest for the 0.5% difference threshold.

**Table 2:** Alignment, major misalignment, and misalignment by observed trend week, threshold and season. This table shows the number and percentage of jurisdiction-weeks each season and overall, that were aligned, major misaligned, or misaligned for each threshold. Major misalignments occurred when *R*_t_ epidemic trend and observed trend indicated changes in opposite directions for a single jurisdiction-week (e.g. growing *R*_t_ epidemic trend category and decreased observed ED trend); misalignments occurred when one metric indicated a change and the other did not (e.g. growing *R*_t_ epidemic trend and stable observed ED trend). Observed thresholds are based on differences between week *w* and week *w + 1*.

| **Threshold** | **Season** | **Total jurisdiction-weeks** | **Aligned** | **Misaligned** | **Major misaligned** |
|---|---|---|---|---|---|
| **0.1% difference** | 2024/2025 | 1472 | 839 (57.0%) | 532 (36.1%) | 101 (6.9%) |
|  | 2025/2026 | 1504 | 808 (53.7%) | 594 (39.5%) | 102 (6.8%) |
|  | **Total** | 2976 | 1647 (55.3%) | 1126 (37.8%) | 203 (6.8%) |
| **0.5% difference** | 2024/2025 | 1472 | 594 (40.4%) | 814 (55.3%) | 64 (4.3%) |
|  | 2025/2026 | 1504 | 542 (36.0%) | 917 (61.0%) | 45 (3.0%) |
|  | **Total** | 2976 | 1136 (38.2%) | 1731 (58.2%) | 109 (3.7%) |
| **5% relative change** | 2024/2025 | 1472 | 922 (62.6%) | 433 (29.4%) | 117 (7.9%) |
|  | 2025/2026 | 1504 | 893 (59.4%) | 492 (32.7%) | 119 (7.9%) |
|  | **Total** | 2976 | 1815 (61.0%) | 925 (31.1%) | 236 (7.9%) |

Alignment patterns varied across the season and by jurisdiction [Figure 2 and Supplement Figures 3–5]. Early in each season, misalignment most commonly involved growing *R_t_* trends with stable observed ED trends, while late in each season misalignment most commonly involved declining or not changing *R_t_* trends with stable or decreasing ED trends, respectively. Most alignment across all jurisdictions occurred in weeks leading up to each season peak when influenza ED visits were steadily increasing. The greatest number of major misalignments occurred during MMWR week 2, ending January 7, 2025, in the 2024/2025 season and during MMWR week 52, ending December 27, 2025, in the 2025/2026 season — both coinciding with epidemic inflection points.

**Figure 2:**
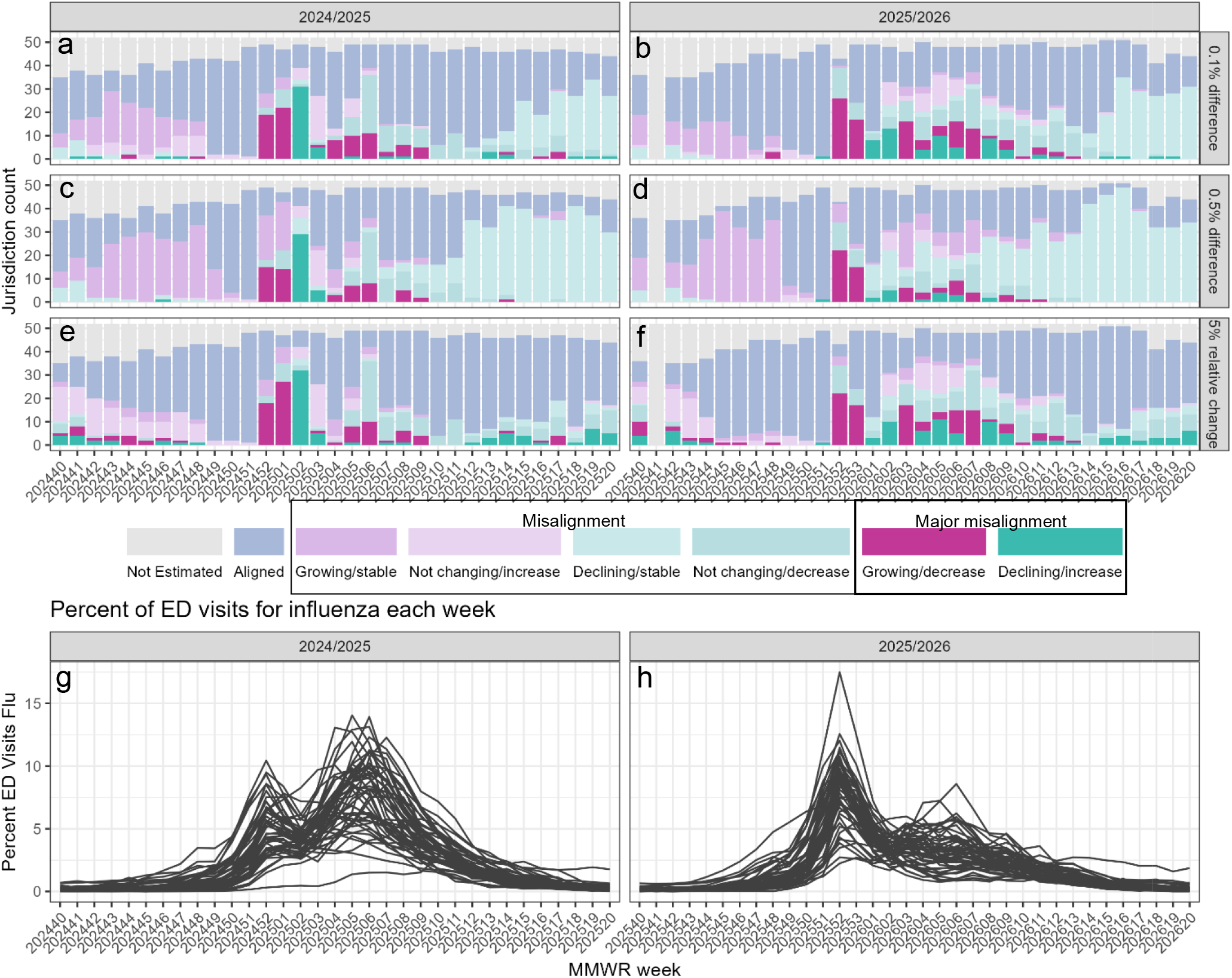
Alignment of *R*_t_ epidemic trend and observed ED trends (panels a–f) and percentage of ED visits for influenza (panels g–h) by week for the 2024/2025 and 2025/2026 seasons. Light blue bars indicate alignment between *R_t_* epidemic trend and observed trend; gray indicates jurisdiction-weeks that were not estimated. Bars are colored by misalignment, with the legend listing the *R_t_* epidemic trend/observed ED trend categories. Darker colored bars indicate a major misalignment (e.g. growing *R_t_* epidemic trend and observed ED trend of decrease) and lighter bars indicate misalignment (e.g. growing epidemic trend and observed ED trend of stable). Each gray line on the bottom graphs represents the percentage of ED visits for influenza for a single jurisdiction by week. Observed thresholds are based on differences between week *w* and week *w + 1*.

Calibration between the probability that *R*_t_ was growing and observed increases in influenza ED visits varied by threshold (Figure 3). The 5% relative change threshold tracked closest to the diagonal, indicating that the predicted probability of growth was reasonably well-calibrated against this definition of observed increase. Calibration was poorer for the two absolute difference thresholds with the 0.5% difference threshold showing the largest deviation from perfect calibration. The number of jurisdiction-weeks per bin by season is available in Supplement Table 1.

**Figure 3:**
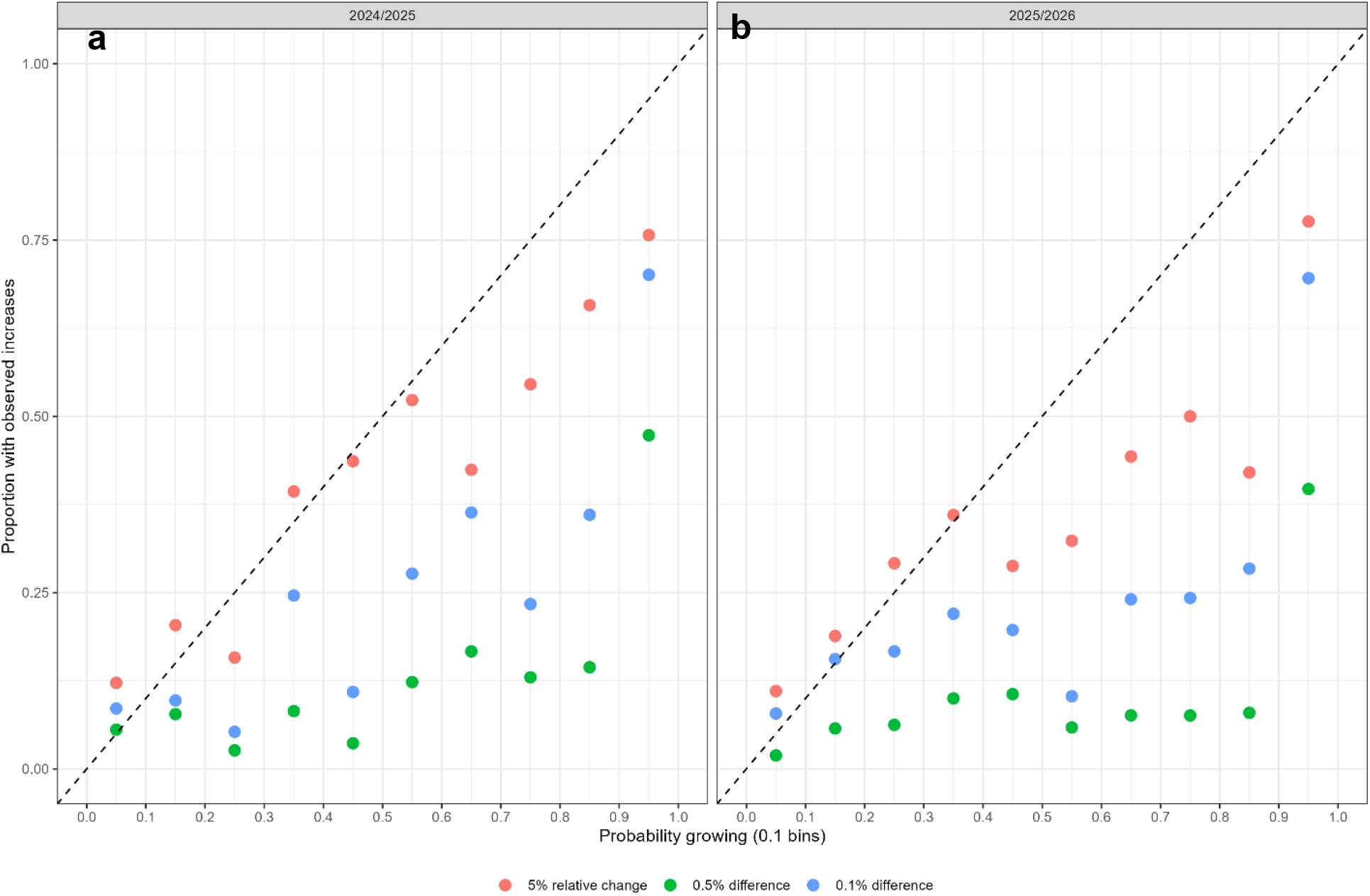
Calibration of the binned probability of increasing infections (probability growing) by the proportion of jurisdiction-weeks with observed increases in influenza ED visits. The probability of increasing infections was binned into 0.1-wide *R*_t_ intervals (x-axis), and the observed proportion of jurisdiction-weeks with an observed increase in influenza ED visits is shown on the y-axis by threshold for the 2024/2025 (panel a) and 2025/2026 (panel b) seasons. The dashed diagonal line represents perfect calibration, where the observed proportion of increases matches the predicted probability of growth. Observed changes and differences are between week *w* and week *w + 1*.

Consistent with the expectation that the probability that *R*_t_ was growing reflects confidence in the direction of the epidemic trend rather than the magnitude of change, the relationship between this probability and the two continuous ED visit-based metrics showed considerable spread across all epidemic trend categories (Figure 4). Jurisdiction weeks classified as growing or declining – reflecting high confidence in the direction of *R*_t_ spanned a wide range of observed relative changes and absolute differences. However, jurisdiction-weeks classified as not changing, where predicted probability was closest to 0.5, also showed more spread in observed changes than anticipated; while we expected observed changes to be small in magnitude in this range given that R_t_ estimates near 1 imply a growth rate close to zero, a subset of these jurisdiction-weeks showed comparatively large relative changes or absolute differences, particularly for 2025/2026.

**Figure 4:**
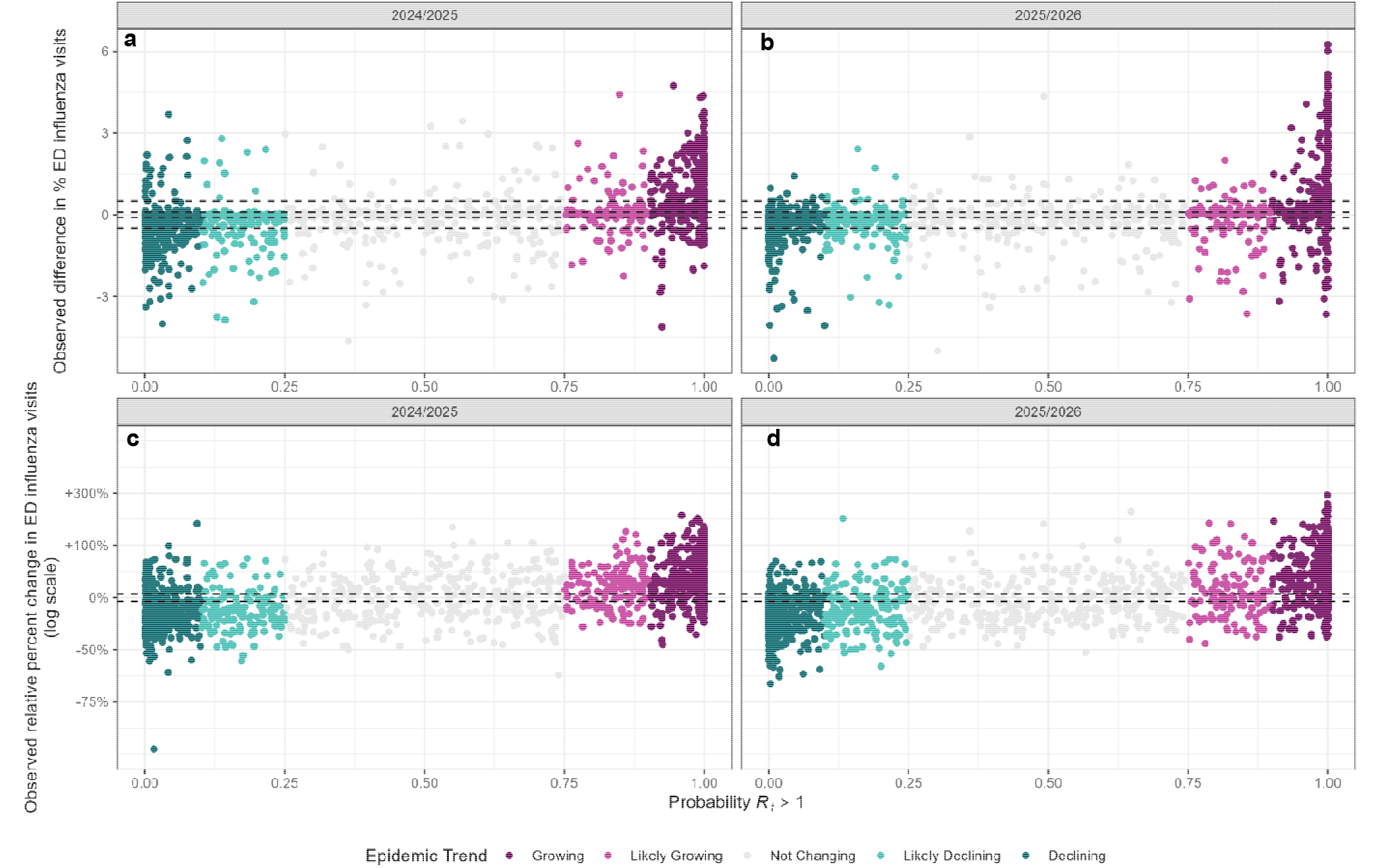
Relationship between the probability that *R_t_*was growing and two observed ED visit-based metrics for each jurisdiction-week: the week-to-week difference in percentage of influenza ED visits (panels a and b) and relative percent change in number of influenza ED visits (panels c and d) for the 2024/25 and 2025/2026 influenza seasons. Each point represents a jurisdiction-week, colored by *R*_t_ epidemic trend categories, which were derived from the probability that *R_t_*was growing. The dashed lines show the 0.5% and 0.1% difference (panels a and b) or the 5% (panels c and d) relative change ED thresholds that were used to determine observed ED trends. Points above the top dashed line would be considered an increase and below the bottom line would be considered a decrease, respectively for each threshold, while points between the dashed lines would be considered stable. Observed changes and differences are between week and week. In panels c and d, relative percent change is plotted on a log scale. Jurisdiction-weeks in which influenza ED visits fell to zero (−100% change; N = 7) cannot be shown on a log scale and are omitted from panels c and d.

From the sensitivity analysis comparing *R*_t_ epidemic trends to concurrent observed ED trends — that is, the difference between week *w* and week *w — 1* rather than week *w* and week *w + 1* — alignment was higher and major misalignment was lower across all thresholds [Supplement Table 2 and Supplement Table 3]. The supplement also includes analogous calibration and relationship figures [Supplement Figures 6-7] that are similar for all thresholds.

## Discussion

We evaluated *R*_t_ epidemic trend categories as early indicators of influenza activity by assessing their alignment with subsequent observed trends in influenza ED visits. We found that alignment varied depending on how changes in ED activity were defined, reflecting important differences between direction-based *R*_t_ epidemic trend categories and magnitude-based measures of observed ED activity. A reasonable applied public health interpretation is that if *R_t_* epidemic trends indicate growing influenza infections, a corresponding increase in influenza ED visits would be observed within a reasonable timeframe. If epidemic trend categories were defined based on estimated *R*_t_ values, not based on P(*R*_t_ >1), they would have a direct relationship to the epidemic growth rate [12] and should therefore correlate strongly with subsequent observed rates of change in ED visits. However, the current practice of defining increasing trend based on the probability *R*_t_ is greater than one muddies these relationships; the expected relationship between the probability infections are increasing and the subsequent rate or size of change in ED visits is not clear. *R_t_* epidemic trend categories showed moderate alignment with observed changes in influenza ED activity, with alignment patterns and diagnostic performance varying considerably across thresholds. Although *R_t_* epidemic trends did not always reliably anticipate changes in ED visits one week ahead, they characterized concurrent trends well and offered a statistically informed complement to simple comparisons of ED visit percentages.

Overall alignment and calibration were strongest for the 5% relative change threshold and weakest for the 0.5% difference threshold. Across all thresholds, the majority of misalignments occurred during major epidemic inflection points. Major misalignments, in particular, likely reflect the fact that near an inflection point *R*_t_ is close to 1, where posterior uncertainty about the direction of change is greatest and small estimation errors are most likely to flip the predicted direction. The expected delay between infection and ED presentation could possibly contribute further to misalignment at these transition points because a rapidly changing growth rate at the time of estimation may no longer reflect the epidemic’s trajectory by the time it is observed in ED visits. Part of this misalignment may be lag-driven, consistent with the sensitivity analysis, in which comparing *R*_t_ epidemic trends to concurrent rather than following-week ED trends reduced major misalignment across all thresholds. This suggests that at least part of the misalignment observed at inflection points could reflect time between infection and care-seeking, rather than a failure of *R*_t_ to capture the epidemic’s true direction at the time of estimation.

In addition, more misalignments occurred at the beginning of the season when influenza ED visits were low. A minimum of ten influenza-associated ED visits is required to publish *R_t_* estimates [4], while a change of less than one visit would constitute a meaningful change under the 5% relative change definition when influenza-associated ED visits are under 20 (e.g.at 15 visits, a 5% change = 0.75 visits). This is one possible cause for the misalignments and major misalignments at the beginning of each season for the 5% relative change threshold when the minimum of 10 ED visits required to publish is met. For the two difference thresholds (i.e. the 0.1 and 0.5% difference), which detect changes in epidemic size, early and late season misalignments largely occurred during periods where the epidemic was growing at a measurable rate but was below the threshold size. If the goal is to anticipate changes in the magnitude of ED visit incidence, *R*_t_ could be combined with other information to estimate the size of change in ED visits, e.g. from an estimate of the force of infection with appropriate lags, or from forecasted ED visit counts based on *R*_t_ estimates. This is consistent with the underlying renewal model and the calibration results, which showed that the probability *R*_t_ >1 tracked reasonably well against the 5% relative change threshold but was poorly calibrated against the absolute difference thresholds. R_t_ estimates near 1 would typically have credible intervals containing values both above and below 1 and therefore be less certain whether R_t_ >1 or R_t_ <1. Since meaningful absolute-scale changes can occur when R_t_ ∼ 1, this framework would likely not consistently capture those changes. A direction-based probability metric alone is insufficient to anticipate the size of change in ED visits, particularly when that change is measured in absolute rather than relative terms. However, when counts are low and R_t_ ∼ 1, both absolute scale differences and relative changes may be less meaningful for public health decision making. Redefining epidemic trend categories using estimated *R*_t_ magnitude, or supplementing them with size-based metrics, could address this gap directly.

Results from the sensitivity analysis comparing *R_t_* epidemic trends to present observed ED trends (i.e. trends observed at the time the *R_t_*estimate was generated rather than the following week) showed consistently stronger alignment across all thresholds. This suggests that *R_t_* epidemic trend categories may perform better as a concurrent indicator of influenza ED activity than as an early one. However, because publicly available trends are delayed, decision-makers may not be able utilize this information as well during the influenza season[14], especially given that observed trends used finalized data and *R*_t_ epidemic trends were generated in real-time with incomplete data. Nevertheless, this does not diminish the value of *R_t_* epidemic trend categories as a surveillance tool as they performed reasonably well in anticipating increases or decreases in influenza ED activity.

In this analysis we evaluated three existing thresholds to assess if influenza emergency department visits are increasing, decreasing, or staying the same. The evaluated thresholds identified different levels of change as meaningful. For example, the 5% relative change threshold first identified influenza increases. For public health practitioners, indications of an early increase could help inform when to implement early season prevention and control strategies, such as encouraging people to get vaccinated before substantial influenza activity is occurring or reminding people to stay home when they are sick. However, the 5% relative change metric also resulted in the fewest weeks classified as stable, potentially making it more difficult to distinguish smaller vs more substantial increases. Conversely, 0.5% absolute difference identifies large changes in the observed burden of influenza ED visits, which may be useful for healthcare surge preparation and response. However, this metric results in the largest number of weeks classified as stable, which could fail to identify smaller, earlier increases in influenza activity. These differences support using different thresholds for different objectives, and this analysis shows that observed influenza trend accuracy depends on the threshold being used.

Several limitations should be considered when interpreting these findings. First, *R*_t_ estimates reflect inferred changes in underlying infections rather than direct measures of ED utilization and increases in infections may not always translate to proportional increases in ED visits due to variation in care-seeking behavior, illness severity, and healthcare system factors. Additionally, *R*_t_ epidemic trend categories represent confidence of an increase whereas observed trend categories indicate magnitude of change. This is evident by the misaligned weeks likely when *R_t_* was truly uncertain about which direction the epidemic was trending and increases and decreases were observed in ED activity compared to when it was confident that the epidemic trend was not changing. Second, differences between the NSSP datasets used for *R*_t_ estimation and those used to define ED observed trend categories may introduce misalignment that reflects data structure rather than true epidemiologic discordance. We also used a final dataset that included backfilled and revised data; one reason for using finalized data was to determine if trend categories were able to capture eventually observed trends, not just artifacts of reporting. Third, this analysis primarily focused on national and state-level results across two influenza seasons, which may not capture the full range of influenza epidemic dynamics across seasons with different onset timing, duration, or severity profiles.

In conclusion, while there was moderate alignment between *R*_t_ epidemic trend categories and observed ED trend categories, these alignments and misalignments should be interpreted in the context of what they measure. *R*_t_ epidemic trend categories reflect confidence in the direction of transmission rather than magnitude, while observed ED trends measure magnitude of change in ED visit counts or percentages. Redefining *R*_t_ epidemic trend categories based on the estimated magnitude of *R_t_*, which has a direct mathematical relationship to the expected growth of ED visits, could improve correspondence with size-based surveillance metrics. Relative change metrics that are more stable at low counts and size-based metrics such as force of infection or forecasted change in ED visits may also be useful directions for future work.

## Data Availability

All data produced in the present study are available upon reasonable request to the authors.

