## Supplemental Figures and Tables for "Utility of *R*_t_ epidemic trend categories for influenza surveillance: alignment with observed emergency department trends across two influenza seasons in the United States"

**Supplement Figure 1:** Observed data and Rt estimate example calendar

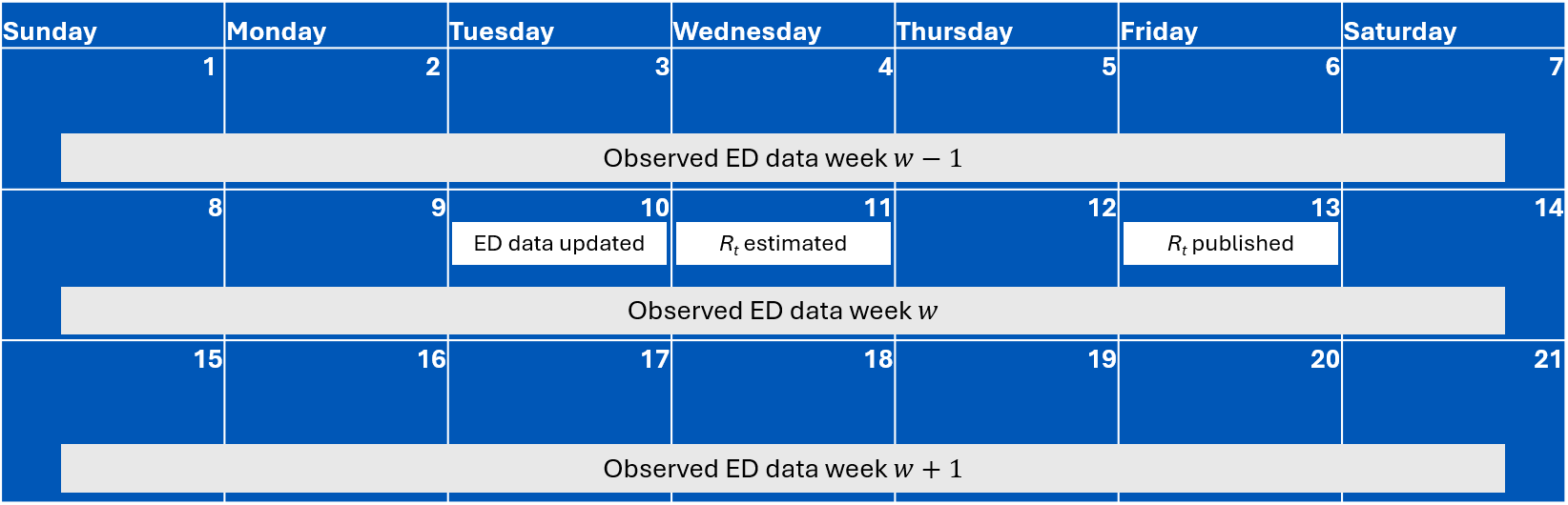

**Supplement Figure 2:** Distribution of weekly absolute (a) and relative (b) differences in emergency department visits across both seasons. The absolute differences (a) are colored by the classification of absolute change at the 0.1% difference threshold, the darker dashed line also indicates the 0.1% threshold, and the lighter dashed line indicates of the 0.5% threshold. The relative differences (b) are colored by the classification of the relative change at the 5% relative change threshold.
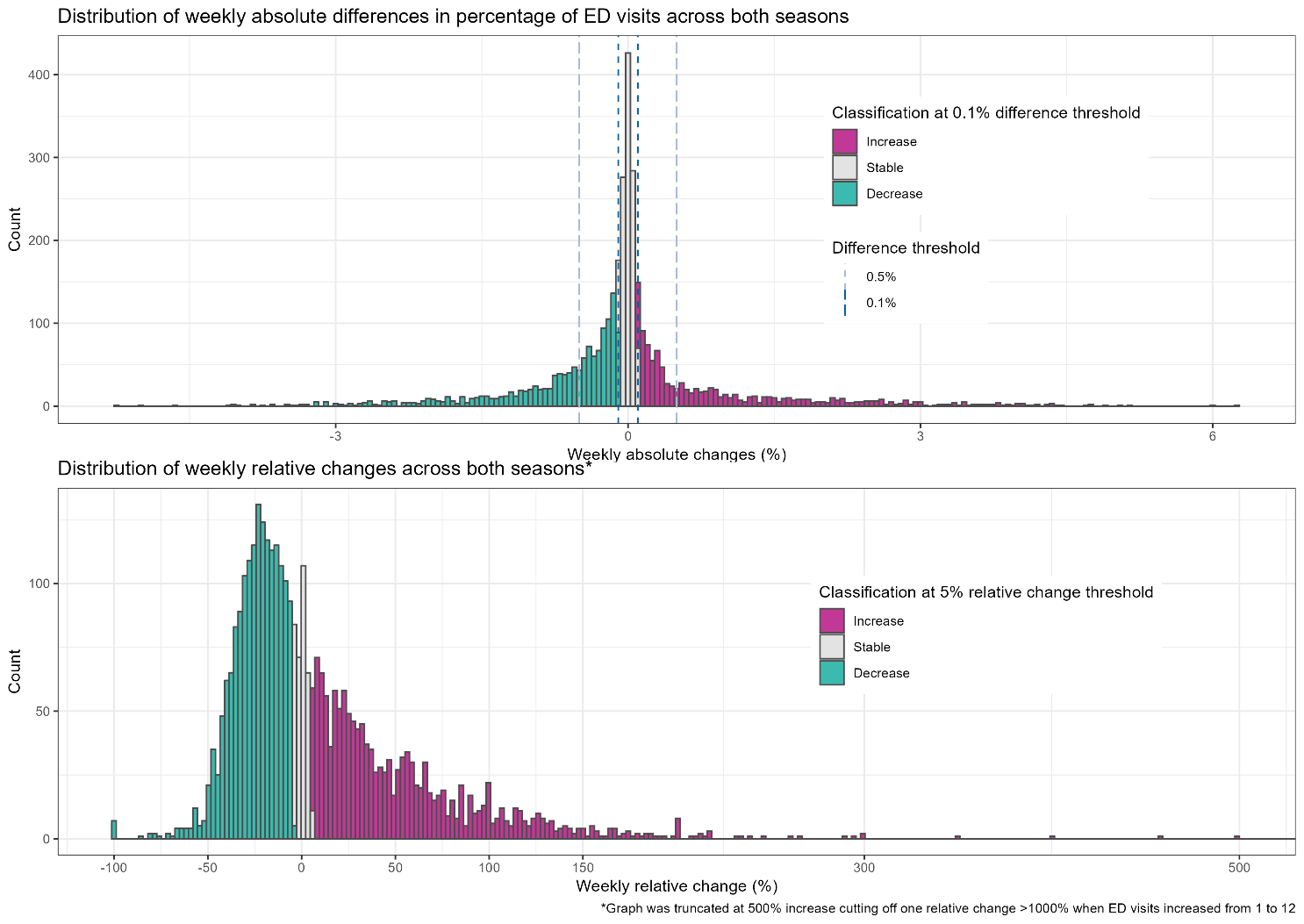

**b**

**a**

**Supplement Figure 3:** Number of weeks aligned by state at the 0.5% difference threshold observed trends.

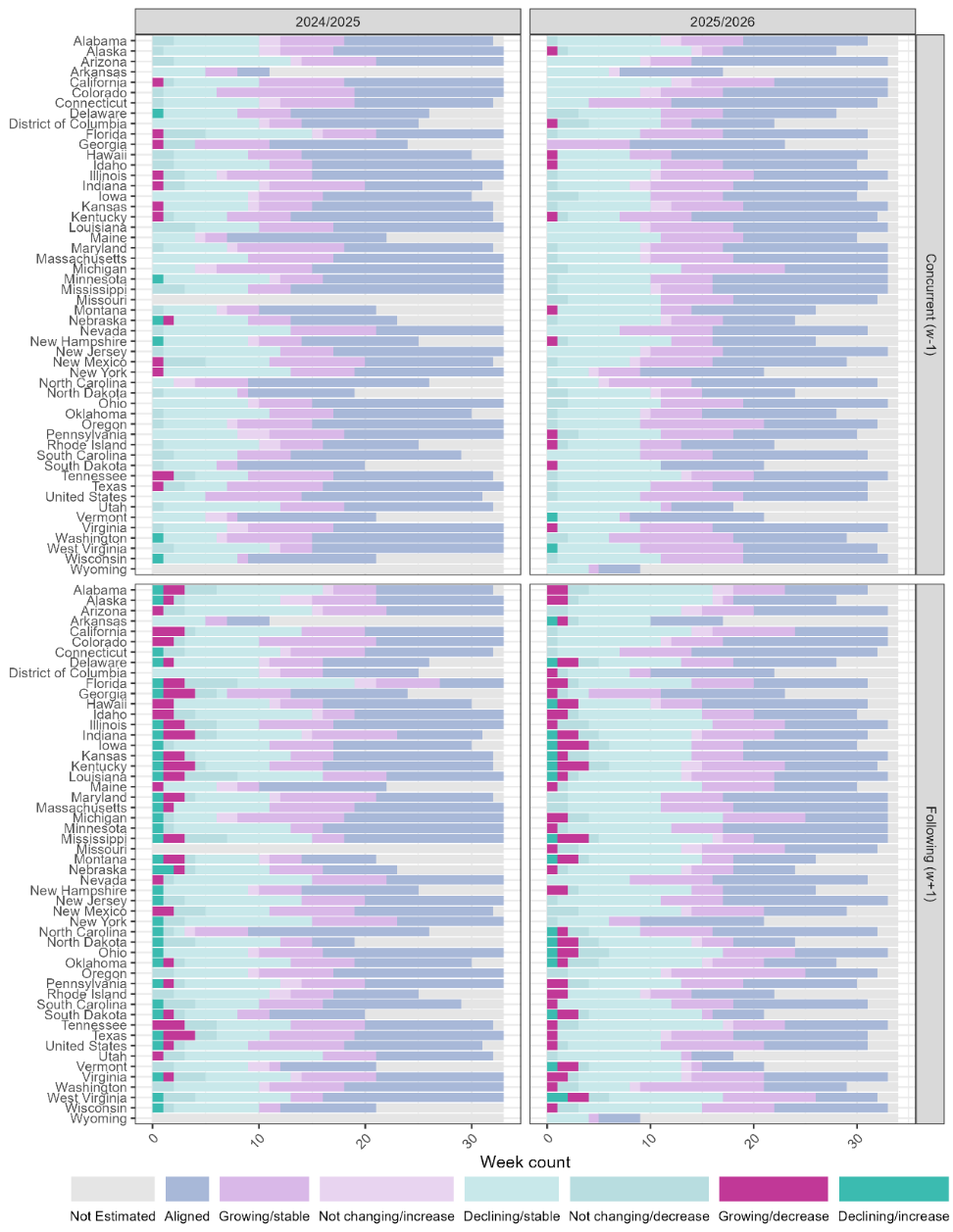

**Supplement Figure 4**: Number of weeks aligned by state at the 0.1% difference threshold observed trends.

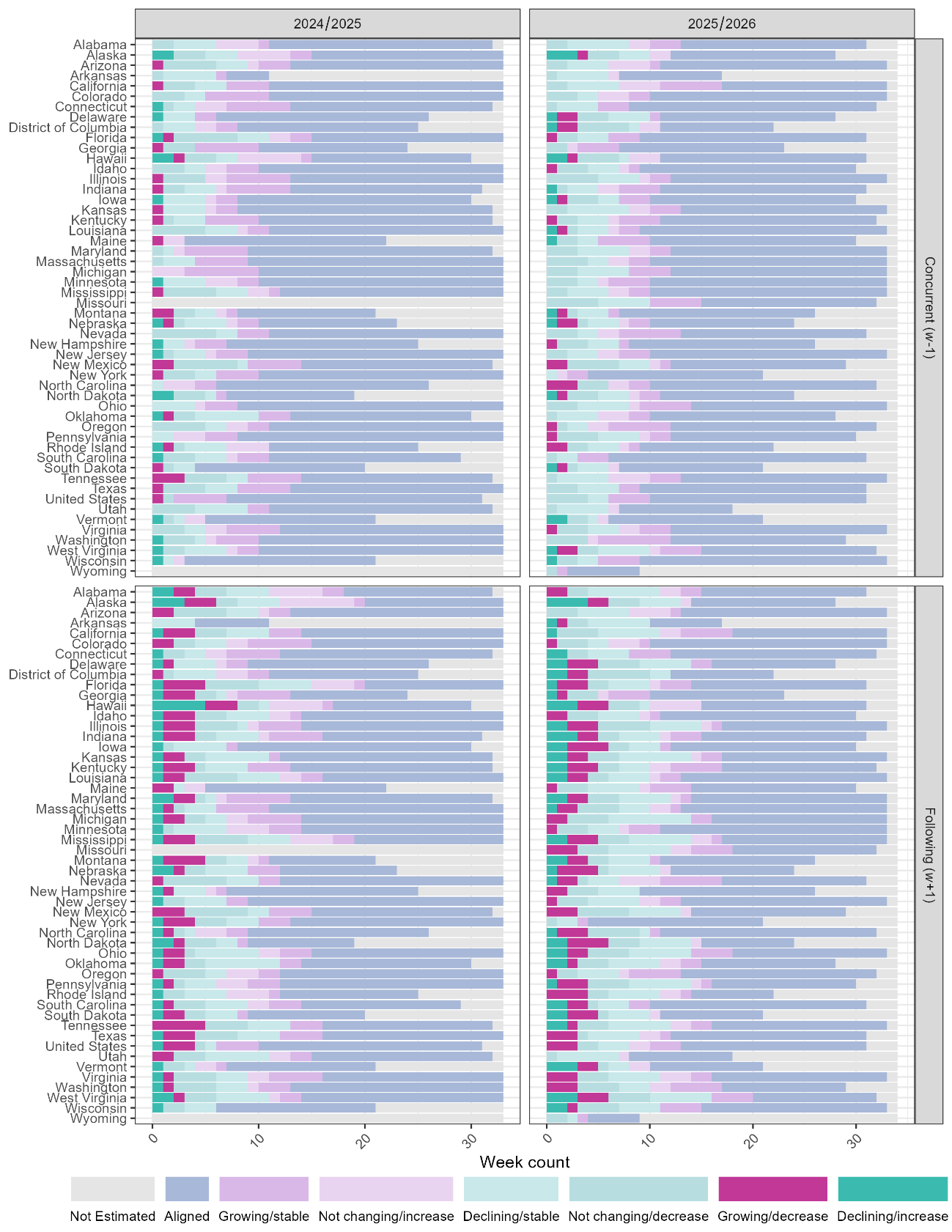

**Supplement Figure 5**: Number of weeks aligned by state at the 5% relative change threshold observed trends.

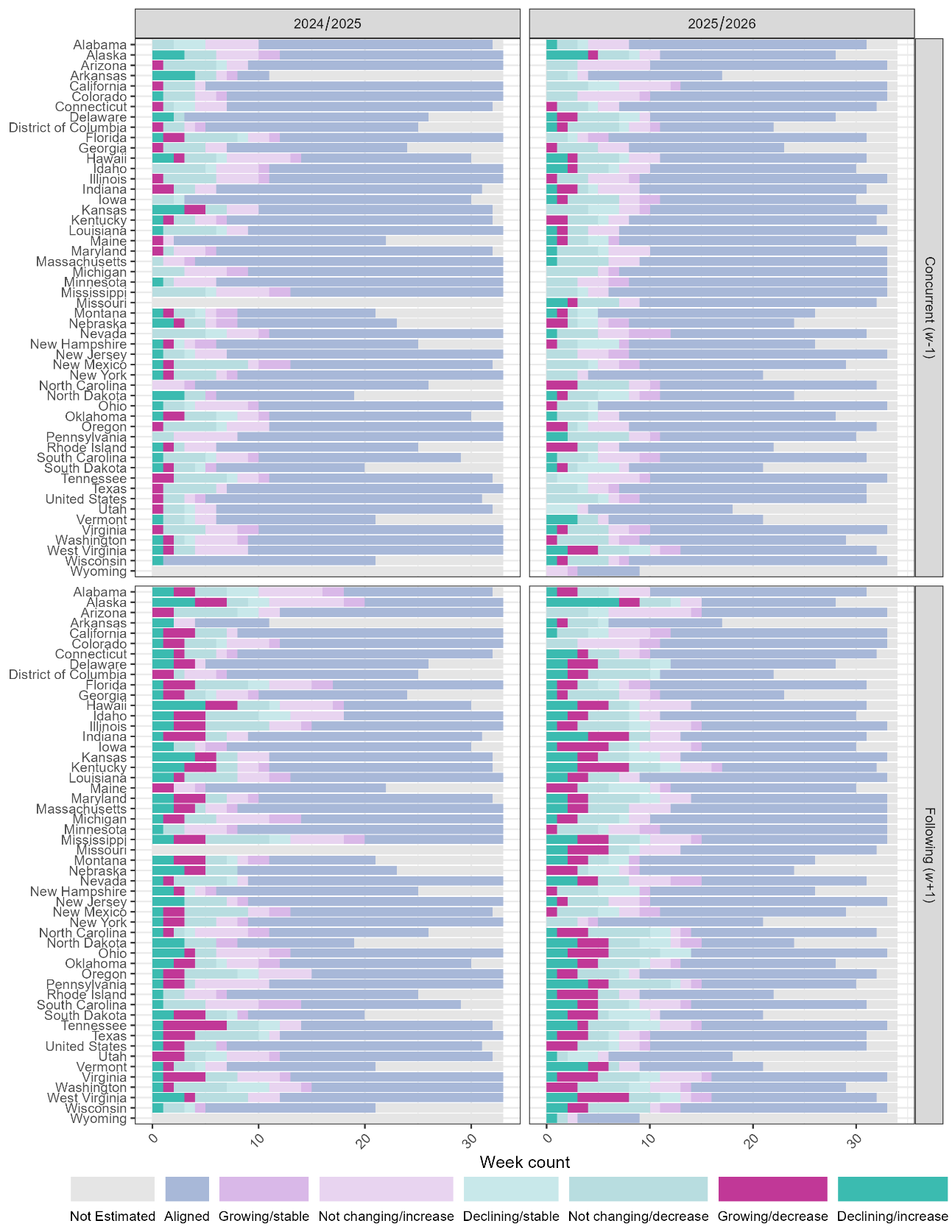

**Supplement Table 1: Number of jurisdiction-weeks per binned probability of increasing *R*_t_ by season.** Probability that *R*_t_ was increasing was binned using 0.1-width bins.

| **Bin** | **2024-2025** | **2025-2026** |
| --- | --- | --- |
| **[0,0.1]** | 467 | 471 |
| **(0.1,0.2]** | 103 | 122 |
| **(0.2,0.3]** | 76 | 96 |
| **(0.3,0.4]** | 61 | 50 |
| **(0.4,0.5]** | 55 | 66 |
| **(0.5,0.6]** | 65 | 68 |
| **(0.6,0.7]** | 66 | 79 |
| **(0.7,0.8]** | 77 | 66 |
| **(0.8,0.9]** | 111 | 88 |
| **(0.9,1]** | 391 | 398 |

**Supplement Table 2**: Alignment, major misalignment, and misalignment by observed trend week, threshold and season. This table shows the number and percent of jurisdiction-weeks each season and overall, that were aligned, major misaligned, or misaligned for each threshold. Observed thresholds are based on differences between week $w$ and week $w-1$.

| **Threshold** | **Season** | **Total jurisdiction-weeks** | **Aligned** | **Misaligned** | **Major misaligned** |
| --- | --- | --- | --- | --- | --- |
| **0.1% difference** | 2024/2025 | 1472 | 978 (66.4%) | 470 (31.9%) | 24 (1.6%) |
|  | 2025/2026 | 1504 | 973 (64.7%) | 513 (34.1%) | 18 (1.2%) |
|  | **Total** | 2976 | 1951 (65.6%) | 983 (33%) | 42 (1.4%) |
| **0.5% difference** | 2024/2025 | 1472 | 719 (48.8%) | 745 (50.6%) | 8 (0.5%) |
|  | 2025/2026 | 1504 | 679 (45.1%) | 821 (54.6%) | 4 (0.3%) |
|  | Total | 2976 | 1398 (47%) | 1566 (52.6%) | 12 (0.4%) |
| **5% relative change** | 2024/2025 | 1472 | 1079 (73.3%) | 366 (24.9%) | 27 (1.8%) |
|  | 2025/2026 | 1504 | 1063 (70.7%) | 420 (27.9%) | 21 (1.4%) |
|  | **Total** | 2976 | 2142 (72%) | 786 (26.4%) | 48 (1.6%) |

**Supplement Table 3:** Count and percent of epidemic trend categories (rows) that aligned with concurrent observed trends defined by 0.5% difference threshold, 0.1% difference threshold, and 5% relative change threshold for both 24/25 and 25/26 seasons. Cells with major misalignment are bolded for visibility. Overall percentages are column percents, percentages in Increase, Stable, Decrease columns are row percents. Observed thresholds are based on differences between week $w$ and week $w-1$.

|  | **Overall** | **Increase** | **Stable** | **Decrease** |
| --- | --- | --- | --- | --- |
| **Threshold 0.1 % difference** | n = 3484 | n = 1002 | n = 1153 | n = 1329 |
| Growing | 789 (22.6%) | 628 (79.6%) | 139 (17.6%) | **22 (2.8%)** |
| Likely Growing | 270 (7.7%) | 95 (35.2%) | 143 (53.0%) | **32 (11.9%)** |
| Not Changing | 650 (18.7%) | 133 (20.5%) | 282 (43.4%) | 235 (36.2%) |
| Likely Declining | 329 (9.4%) | **20 (6.1%)** | 101 (30.7%) | 208 (63.2%) |
| Declining | 938 (26.9%) | **20 (2.1%)** | 180 (19.2%) | 738 (78.7%) |
| Not Estimated | 508 (14.6%) | 106 (20.9%) | 308 (60.6%) | 94 (18.5%) |
| **Threshold 0.5% difference** | n = 3484 | n = 531 | n = 2317 | n = 636 |
| Growing | 789 (22.6%) | 382 (48.4%) | 396 (50.2%) | **11 (1.4%)** |
| Likely Growing | 270 (7.7%) | 41 (15.2%) | 216 (80.0%) | **13 (4.8%)** |
| Not Changing | 650 (18.7%) | 61 (9.4%) | 492 (75.7%) | 97 (14.9%) |
| Likely Declining | 329 (9.4%) | **7 (2.1%)** | 215 (65.3%) | 107 (32.5%) |
| Declining | 938 (26.9%) | **1 (0.1%)** | 561 (59.8%) | 376 (40.1%) |
| Not Estimated | 508 (14.6%) | 39 (7.7%) | 437 (86%) | 32 (6.3%) |
| **Threshold 5% relative change** | n = 3483 | n = 1459 | n = 337 | n = 1687 |
| Growing | 789 (22.7%) | 741 (93.9%) | 29 (3.7%) | **19 (2.4%)** |
| Likely Growing | 270 (7.8%) | 166 (61.5%) | 51 (18.9%) | **53 (19.6%)** |
| Not Changing | 650 (18.7%) | 239 (36.8%) | 119 (18.3%) | 292 (44.9%) |
| Likely Declining | 329 (9.4%) | **43 (13.1%)** | 29 (8.8%) | 257 (78.1%) |
| Declining | 938 (26.9%) | **29 (3.1%)** | 50 (5.3%) | 859 (91.6%) |
| Not Estimated | 507 (14.6%) | 241 (47.5%) | 59 (11.6%) | 207 (40.8%) |

**Supplement Figure 6: Calibration of the binned probability of increasing infections by observed increases in influenza ED visits.** The probability that *R_t_* was growing was binned into 0.1-wide *R_t_* intervals (x-axis), and the observed proportion of jurisdiction-weeks with an observed increase in influenza ED visits is shown on the y-axis by threshold. The dashed diagonal line represents perfect calibration, where the observed proportion of increases matches the predicted probability of growth**.** Observed changes and differences are between week $w$ and week $w-1$.

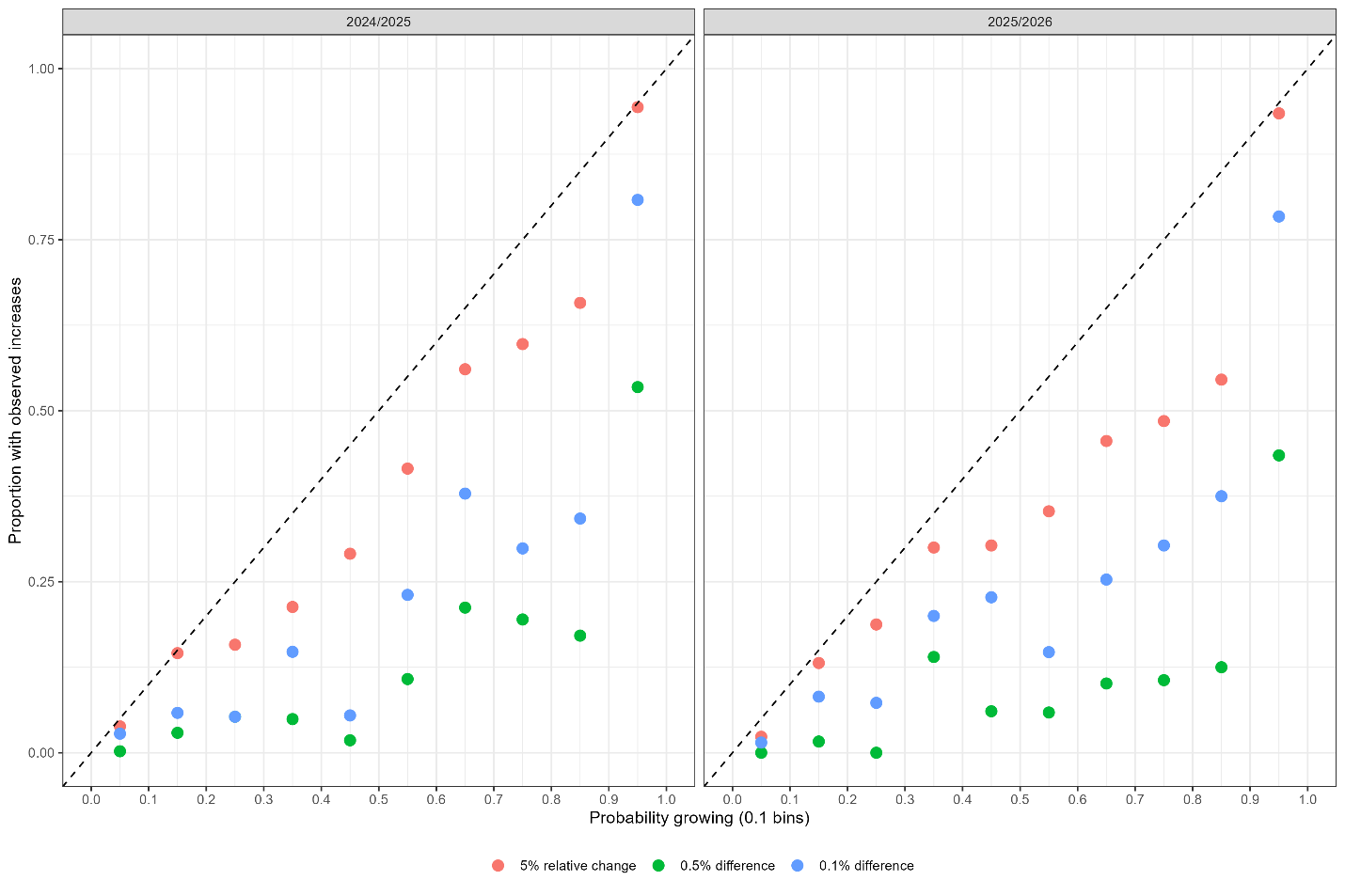

**Supplement Figure 7**: **Probability influenza epidemic is growing by concurrent observed percent change of influenza ED visits and by concurrent relative change in percent of influenza ED visits.** Points are colored by epidemic trend and the dashed lines show the 0.5% and 0.1% thresholds of change or the 5% relative change thresholds that were used in determining increases and decreases for observed concurrent trends. Points above the dashed line would be considered increases and decreases, respectively for each threshold, while points between the dashed lines would be considered stable. Observed thresholds are based on differences between week $w$ and week $w-1$.

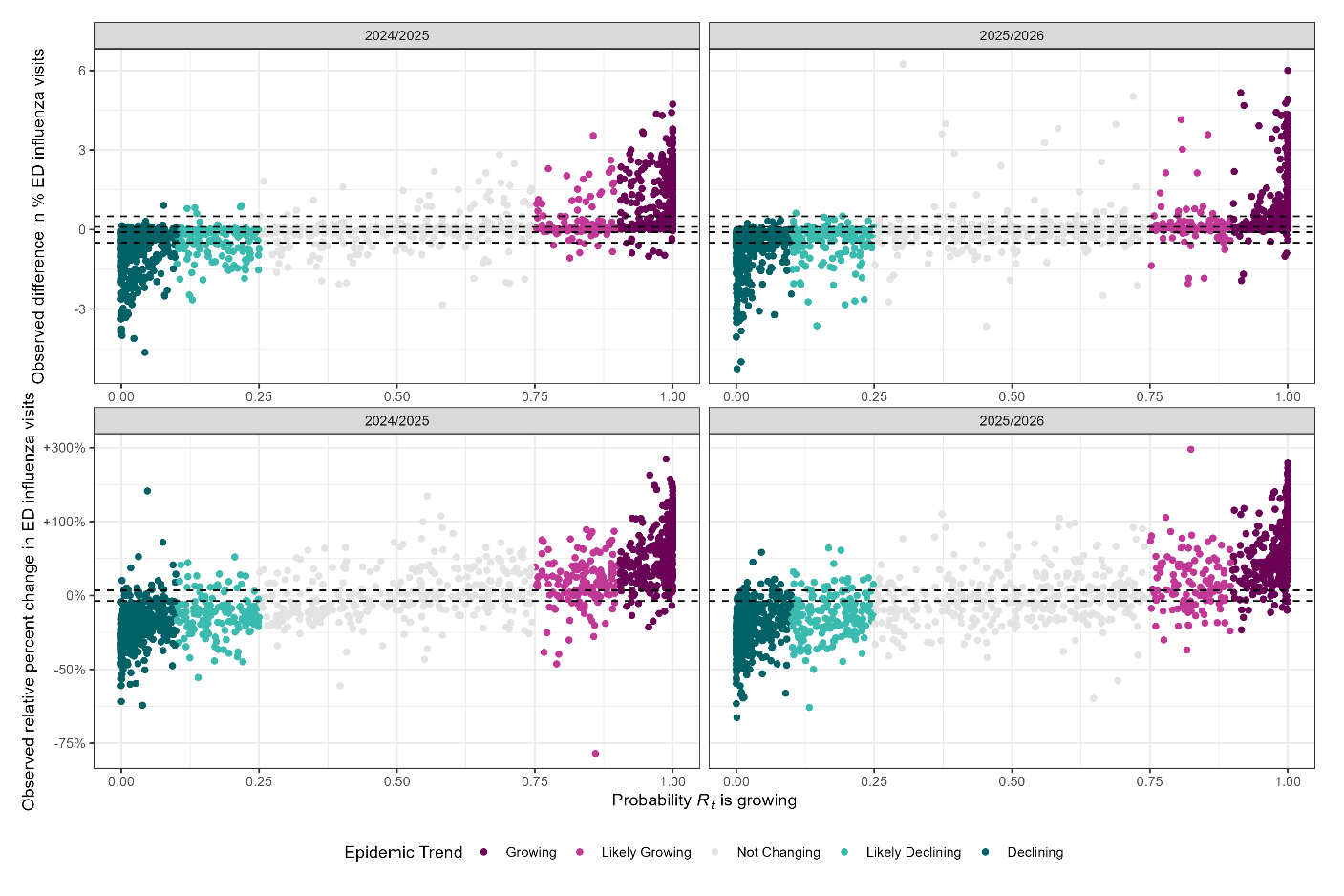
